# Epidemiological analysis of all varicose vein surgeries over 9 years in Brazil: trends, costs and mortality rate of 1,266,550 cases

**DOI:** 10.1101/2025.03.04.25323381

**Authors:** Alexandre Fioranelli, Bruno Jeronimo Ponte, Carolina Carvalho Jansen Sorbello, Felipe Soares Oliveira Portela, Andressa Cristina Sposato Louzada, Marcelo Fiorelli Alexandrino da Silva, Marcelo Passos Teivelis, Nelson Wolosker

## Abstract

2

**Objectives:** To evaluate the total number of surgical procedures for varicose vein treatment performed within the Brazilian Unified Public Health System (SUS) and the Private Health System (PHS) between 2015 and 2023 and to analyze their differences regarding the number of procedures over time, financial investments, and in-hospital mortality rates.

**Materials and Methods:** Data on varicose vein surgeries performed between 2015 and 2023 were extracted from the SUS database and the procedural code table provided by the National Supplementary Health Agency (ANS), which reports procedures conducted within the PHS in Brazil. The data included geographic region, number of procedures performed, in-hospital mortality (reported only by SUS), and the amount invested by the government and private health insurance providers.

**Results:** A total of 1,266,550 varicose vein surgeries were performed in both public and supplementary healthcare systems in Brazil between 2015 and 2023, with an average surgery rate per 10,000 inhabitants per year of 3.7 in SUS and 14.8 in PHS. The total financial investment amounted to BRL 1,492,310,372.13, with an overall average cost per procedure of BRL 1,240.92. The mean cost per procedure in SUS was BRL 679.50, whereas in PHS, it was BRL 1,632.60—approximately 3.15 times higher than in SUS. A total of 29 in-hospital deaths were recorded in SUS after varicose vein surgery, representing an overall mortality rate of 0.005%.

**Conclusions:** A total of 1,266,550 surgeries were performed between 2015 and 2023 for chronic venous disease treatment, with a rate of 6.5 procedures per 10,000 inhabitants per year. In the PHS system, the number of surgeries was four times higher than in SUS, considering the number of procedures per 10,000 inhabitants. The total investment in PHS was 3.15 times greater than in SUS, with an estimated average cost per procedure of BRL 1,632.60. The observed in-hospital mortality rate in SUS was 0.005%.

## 4. Introduction

Chronic Venous Disease (CVD) is a common and progressive condition that poses a significant public health challenge (1,2). It affects the quality of life for many individuals and generates substantial medical costs. (3,4) It is estimated that over 50% of the population may be affected by CVD, particularly older adults. The disease is associated with worsening outcomes, including venous ulcer development, which can lead to increased morbidity and socioeconomic productivity losses. (5–7)

Complications of CVD include deep vein thrombosis (DVT) (1) and variceal hemorrhage, both of which can be life-threatening. (8,9) The economic impact is considerable, contributing to productivity loss and early retirement. In the United States, annual costs exceed $3 billion, highlighting this disease’s financial and social burden. (10)

Surgical treatment options for CVD include traditional methods (ligation and stripping) and minimally invasive techniques, which have shown high efficacy and cost-effectiveness. (11–13) Despite their overall safety, some complications can arise, including hemorrhage, deep vein thrombosis, and pulmonary embolism. (13–15) In developed countries, the 30-day postoperative mortality rate is low, estimated at less than 0.02%, according to national registries. (16–18)

In Brazil, a study conducted by Silva et al. analyzed data from the Unified Public Health System (SUS) (public healthcare system) from 2008 to 2019, documenting 869,220 procedures for CVD treatment. (19) The mortality rate was 0.0056%, and the total costs exceeded $232 million. Currently, approximately 75% of the Brazilian population relies exclusively on SUS, while 25% primarily access the private healthcare sector. (19)

At the time of Silva et al.’s publication, data on procedures performed in the private healthcare sector (PHS) were unavailable, resulting in the exclusion of 25% of the population from the analysis. Consequently, real-world data that includes the entire Brazilian population and comparative studies between the public and private healthcare systems for these procedures have not been reported.

However, data on procedures conducted in the PHS have recently become publicly available, covering interventions conducted between 2015 and 2023.

This study aims to retrospectively analyze surgical procedures performed in both the public and private healthcare systems to treat CVD from 2015 to 2023. By examining an estimated population of 210 million Brazilians, the study compares the two systems in terms of procedural volume, costs, patient characteristics, and temporal trends. The findings will provide comparative data to support the development of effective strategies for CVD management in Brazil.

## 5. Materials and Methods

All surgical procedures for the treatment of chronic venous insufficiency performed within SUS and the PHS were analyzed.

This research was approved by the institution’s Ethics Committee, which is responsible for its conduction. All data obtained from DATASUS, TabNet, and the PHS (D-TISS) were de-identified, ensuring participant anonymity. As a result, the Institutional Review Board (IRB) granted a waiver for informed consent.

Data were retrieved from TabNet, a public health information tool developed by DATASUS, part of the Department of Informatics and data from SUS under the Brazilian Ministry of Health, as well as from the supplementary healthcare system platform. TabNet provides open-access data on procedures conducted within SUS, which are reported by accredited hospitals and outpatient clinics. Accreditation and data reporting are mandatory for government reimbursement of these procedures.

The supplementary healthcare system platform (D-TISS), is managed by the National Supplementary Health Agency (ANS) and provides open-access data on procedures conducted within this sector.

Data on vascular surgical procedures for the treatment of CVD was collected from the TabNet and D-TISS platforms, covering the period from 2015 to 2023. The collected data included geographic region, the number of procedures performed, in-hospital or outpatient mortality (for SUS only), and the financial investments made by both SUS and PHS.

For data extraction within SUS, two procedure types were evaluated according to the coding system established by the SUS Procedure, Medicine, and OPM Management System (SIGTAP): bilateral surgical treatment for CVD (04.06.02.056-6) and unilateral surgical treatment for CVD (04.06.02.057-4).

Data collection for the SS sector was obtained from the D-TISS Data Panel, published by ANS, and procedures were analyzed according to the PHS coding system, which includes: Surgical treatment of varicose veins with lipodermatosclerosis or ulcer (30907101); Surgical treatment of varicose veins in both limbs (30907136); Surgical treatment of varicose veins in one limb (30907144); Resection of collateral veins under local anesthesia in an outpatient setting (30907152).

Data on annual average and total accumulated inflation between 2015 and 2023 (5.3% and 59.3%, respectively) were obtained from the Brazilian Institute of Geography and Statistics (IBGE) to analyze financial investments in SUS and PHS. (20)

All data were collected from publicly accessible websites using automated web scraping programs. The automation scripts were developed in Python (v. 2.7.13, Beaverton, OR, USA) using the Windows 10 Single Language operating system. Data collection, field selection within the platform, and table adjustments were performed using the selenium-webdriver (v. 3.1.8, Selenium HQ, global contributors) and pandas (v. 2.7.13, Lambda Foundry, Inc. & PyData Development Team, New York, NY, USA) packages. The Mozilla Firefox browser (v. 59.0.2, Mountain View, CA, USA) and geckodriver (v. 0.18.0, Mozilla Corporation, Bournemouth, UK) were used for web navigation.

After collecting and processing the data, all information was organized and compiled into a spreadsheet using Microsoft Office Excel 2016® (v. 16.0.4456.1003, Redmond, WA, USA).

### Statistical Analysis

Linear regression was used to assess trends in the distribution of procedures across Brazilian regions over time, as well as their associated costs. An unpaired Student’s t-test was conducted to compare quantitative variables, including the number of procedures, financial investments, and patient age in SUS and PHS. Categorical variables were analyzed using the Chi-square test to compare distributions between SUS and PHS. Statistical significance was set at p < 0.05.

## 6. Results

Between 2015 and 2023, 1,266,550 surgical procedures were performed to treat CVD in public and private hospitals in Brazil. The number and frequency of CVD surgeries over the years are presented in Table 1. The PHS accounted for 57% of all CVD procedures performed in Brazil during this period. Throughout the years, the PHS consistently performed a higher percentage of surgeries than SUS, except in 2018, when both systems conducted an equal share of 50% of procedures.

**Table 1.** Absolute and relative values of the number of procedures performed to treat varicose veins in the SUS and the PHS between 2015 and 2023.

| Ano | SUS |  | PHS |  | Total |
| --- | --- | --- | --- | --- | --- |
|  | *n* | (%) | *n* | (%) |  |
| **2015** | 69,114 | 44% | 88,162 | 56% | 157,276 |
| **2016** | 68,885 | 47% | 77,980 | 53% | 146,865 |
| **2017** | 67,131 | 44% | 84,617 | 56% | 151,748 |
| **2018** | 75,125 | 50% | 76,148 | 50% | 151,273 |
| **2019** | 68,726 | 44% | 86,499 | 56% | 155,225 |
| **2020** | 28,564 | 37% | 47,912 | 63% | 76,476 |
| **2021** | 25,598 | 30% | 60,509 | 70% | 86,107 |
| **2022** | 55,521 | 38% | 90,901 | 62% | 146,422 |
| **2023** | 68,190 | 44% | 86,968 | 56% | 155,158 |
| **Total** | 526,854 | 43% | 699,696 | 57% | 1,226,550 |

*SUS: Unified public health system; PHS: private health system.*

The distribution of procedure rates per 10,000 inhabitants is illustrated in Figure 1. The rates for public healthcare were calculated based on the population that exclusively relies on the public system (75% of the Brazilian population). In contrast, the rates for the PHS procedures were based on the population primarily using private healthcare (25% of the Brazilian population). Overall rates were estimated using the total Brazilian population, as the Brazilian Institute of Geography and Statistics (IBGE) reported during the analyzed period.(21)

**Figure 1.**
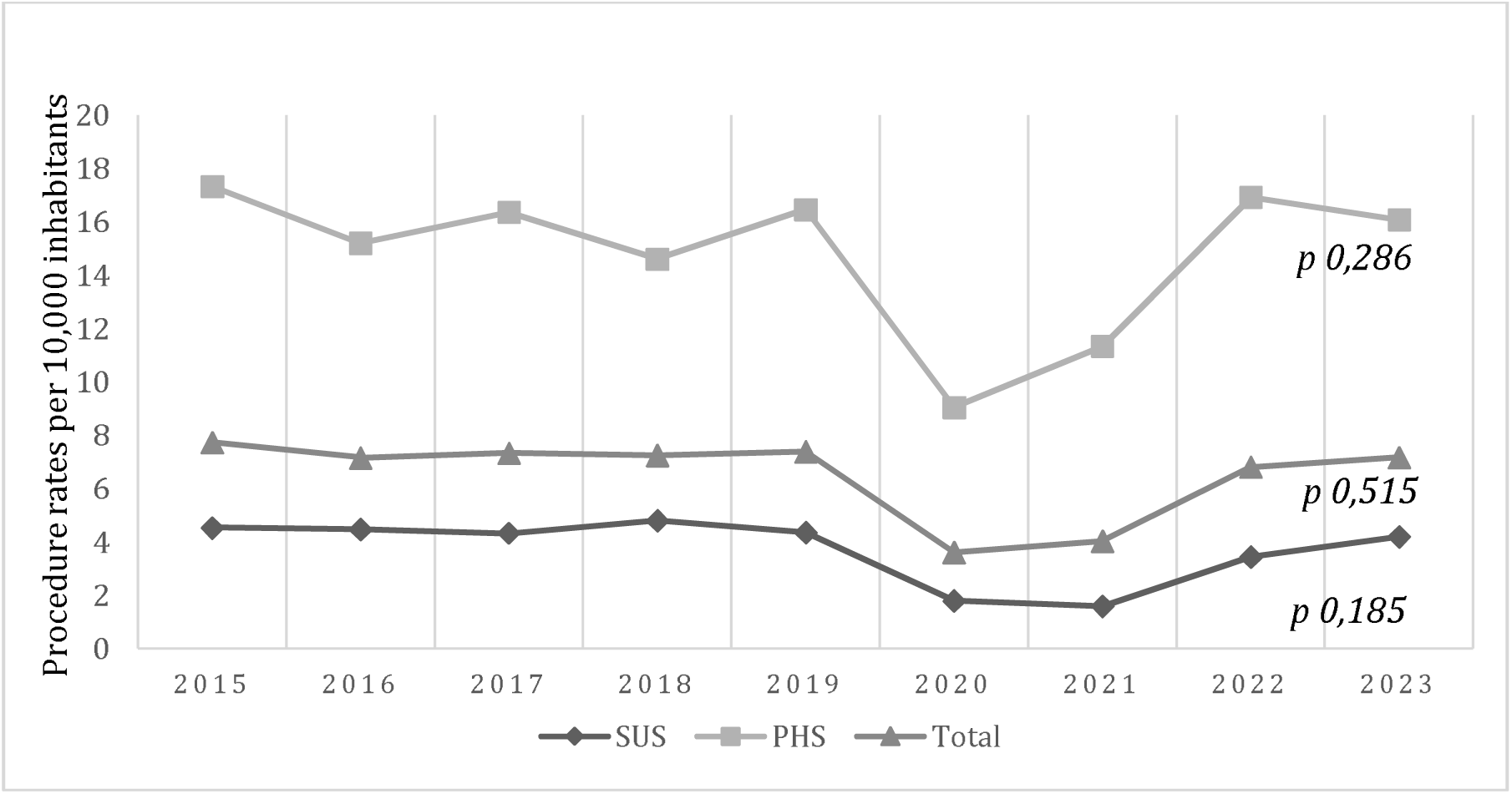
Distribution of procedures performed per 10,000 inhabitants among exclusive SUS users, supplementary health users, and the general population. SUS: unified public health system; PHS: private health system.

The average rate in the public healthcare system was 3.7 CVD surgeries per 10,000 inhabitants annually. In the PHS, this rate was significantly higher, at 14.8 surgeries per 10,000 inhabitants, which is four times greater than of the public system *(p<0.05)*.

A considerable decline in surgeries was noted in 2020 and 2021. Compared to 2019, there was a decrease of 58.75%, 45%, and 51.10% in the number of CVD procedures performed in the public, private, and overall populations, respectively. There was not a significant trend toward an increase or decrease in the number of CVD procedures. The recorded data from the SUS (*p = 0.185*), PHS (*p = 0.515*), and total (*p = 0.286*) did not demonstrate statistically significant temporal trends.

The total financial investments for each healthcare system are shown in Table 2. The combined expenditures by the government and private health insurers from 2015 to 2023 amounted to R$ 1,492,310,372.13. The public healthcare system allocated R$ 359,539,892.13 for CVD treatment procedures, while private insurers spent R$ 1,132,770,480.00—3.15 times more than the public system *(p<0.05)* (Figure 2).

**Figure 2.**
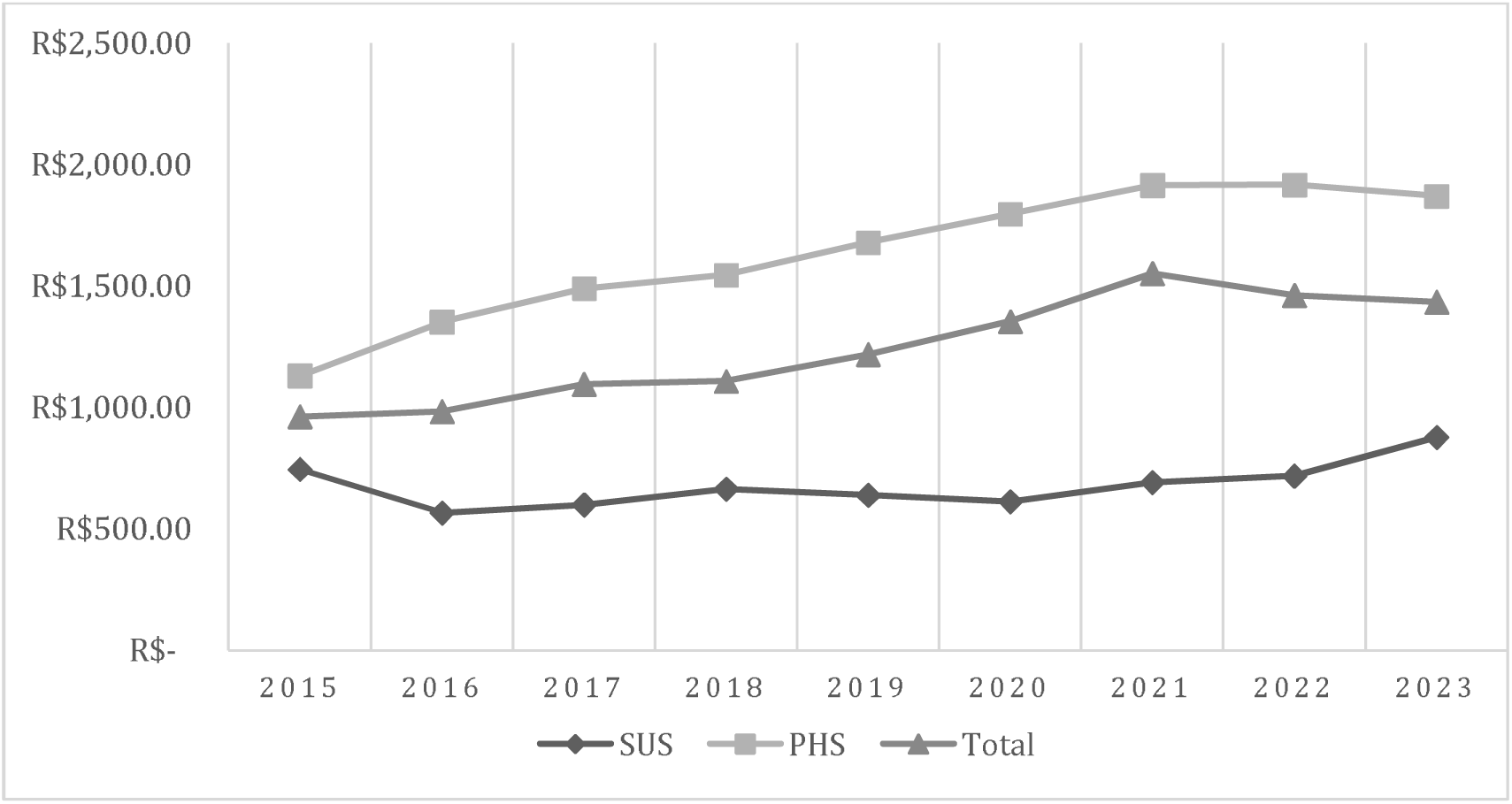
Average transfer per procedure in SUS, supplementary health and total between 2015 and 2023. SUS: Unified public health system; PHS: private health system.

**Table 2.** Investments made in SUS and supplementary health procedures between 2015 and 2023.

| Year | SUS |  |  | PHS |  |  | Total |  |  |
| --- | --- | --- | --- | --- | --- | --- | --- | --- | --- |
|  | Value | (%) | Average | Value | (%) | Average | Value | Average |  |
| 2015 | R\$ 51,511,581.77 | 34% | R\$ 745.31 | R\$ 99,683,770.00 | 66% | R\$ 1,130.69 | R\$ 151,195,351.77 | R\$ 961. | |
| 2016 | R\$ 39,004,929.95 | 27% | R\$ 566.23 | R\$ 105,416,540.00 | 73% | R\$ 1,351.84 | R\$ 144,421,469.95 | R\$ 983. | |
| 2017 | R\$ 40,262,492.52 | 24% | R\$ 599.76 | R\$ 126,048,500.00 | 76% | R\$ 1,489.64 | R\$ 166,310,992.52 | R\$ 1,095. | |
| 2018 | R\$ 49,930,542.78 | 30% | R\$ 664.63 | R\$ 117,684,370.00 | 70% | R\$ 1,545.47 | R\$ 167,614,912.78 | R\$ 1,108. | |
| 2019 | R\$ 43,922,479.20 | 23% | R\$ 639.10 | R\$ 145,207,320.00 | 77% | R\$ 1,678.72 | R\$ 189,129,799.20 | R\$ 1,218. | |
| 2020 | R\$ 17,519,737.59 | 17% | R\$ 613.35 | R\$ 86,045,320.00 | 83% | R\$ 1,795.90 | R\$ 103,565,057.59 | R\$ 1,354. | |
| 2021 | R\$ 17,715,195.56 | 13% | R\$ 692.05 | R\$ 115,879,340.00 | 87% | R\$ 1,915.08 | R\$ 133,594,535.56 | R\$ 1,551. | |
| 2022 | R\$ 39,845,500.76 | 19% | R\$ 717.67 | R\$ 174,198,700.00 | 81% | R\$ 1,916.36 | R\$ 214,044,200.76 | R\$ 1,461. | |
| 2023 | R\$ 59,827,432.00 | 27% | R\$ 877.36 | R\$ 162,606,620.00 | 73% | R\$ 1,869.73 | R\$ 222,434,052.00 | R\$ 1,433. | |
| **Total** | **R\$ 359,539,892.13** | **24%** | **R\$ 679.50** | **R\$ 1,132,770,480.00** | **76%** | **R\$ 1,632.60** | **R\$ 1,492,310,372.13** | **R\$ 1,240** | |

*SUS: Unified public health system; SS: Private health system.*

The average annual expenditure per procedure in Brazil was R$ 1,240.92. In the PHS, the annual cost per procedure was R$ 1,632.60, which was 2.4 times higher than the cost in the public system, which averaged R$ 679.50 *(p<0.05)*.

Annual expenditures for CVD treatment procedures progressively increased over the years. In the public healthcare system, no significant growth was observed, and the per-procedure costs did not adjust adequately for the accumulated inflation rate (2023 value: R$ 877.36 vs. inflation-adjusted value: R$ 1,187.35) *(p = 0.131).* Conversely, PHS expenditures demonstrated linear growth, slightly below the inflation-adjusted value (2023 value: R$ 1,801.30 vs. inflation-adjusted value: R$ 1,876.94) *(p<0.05*), contributing to the overall increase in CVD surgery-related costs in Brazil *(p<0.05)*.

A total of 77.65% of surgeries were performed on female patients. Analysis of both the public and private healthcare systems confirmed that the majority of cases involved female patients in both settings *(p<0.05).* In the public system, 79% of cases were female, while in the PHS, 76% of patients were female. Patient sex distribution is detailed in Table 3.

**Table 3.** Distribution by gender of procedures performed to treat CVD.

| Sexo | SUS |  | PHS |  | Total |  |
| --- | --- | --- | --- | --- | --- | --- |
|  | *n* | (%) | *n* | (%) | *n* | (%) |
| Men | 108.181 | 20,53% | 155.418 | 22,21% | 263.599 | 21,50% |
| Women | 418.673 | 79,47% | 533.802 | 76,29% | 952.475 | 77,65% |
| Not informed | 0 | 0,00% | 10.476 | 1,50% | 10.476 | 0,85% |

*SUS: Unified public health system; PHS: Private health system; CVD: Chronic Venous Disease*

Regarding patient age, the majority of cases occurred in individuals aged 40–49 years (28.15%). In the public healthcare system, most cases were in patients aged 40–59 years (57.25%), whereas in the PHS, procedures were more common in younger patients aged 30–49 years (53.64%) (p<0.05). The age distribution is shown in Table 4.

**Table 4.** Distribution by age group of procedures carried out in the SUS, complementary health and total procedures.

| Age (Years) | SUS |  | PHS |  | Total |  |
| --- | --- | --- | --- | --- | --- | --- |
|  | *n* | % | *n* | % | *n* | % |
| >80 | 1,392 | 0.26% | 2,642 | 0.38% | 4,034 | 0.33% |
| 70-79 | 19,288 | 3.66% | 25,878 | 3.70% | 45,166 | 3.68% |
| 60-69 | 94,611 | 17.96% | 82,738 | 11.82% | 177,349 | 14.46% |
| 50-59 | 155,941 | 29.60% | 151,376 | 21.63% | 307,317 | 25.06% |
| 40-49 | 145,669 | 27.65% | 199,635 | 28.53% | 345,304 | 28.15% |
| 30-39 | 86,711 | 16.46% | 175,662 | 25.11% | 262,373 | 21.39% |
| 20-29 | 21,691 | 4.12% | 48,769 | 6.97% | 70,460 | 5.74% |
| < 20 | 1,550 | 0.29% | 2,347 | 0.34% | 3,897 | 0.32% |
| Not informed | 1 | <0.01% | 10,649 | 1.52% | 10,650 | 0.87% |
| **Total** | 526,854 | 100.00% | 699,696 | 100.00% | 1,226,550 | 100.00% |

*SUS: Unified public health system; PHS: Private health system.*

The regional distribution of procedures across Brazil is presented in Table 5. A concentration of surgeries was observed in the Southeast (57.70%) and South (23.33%) regions. Regions with lower population density, such as the Midwest (6.96%) and North (1.44%), reported the lowest procedure rates. Except for the North, all regions had a higher number of procedures performed within the PHS compared to the public healthcare system. No statistically significant temporal trends were observed in the regional distribution of procedures from 2015 to 2023, as analyzed through linear regression. No statistically significant temporal trends were observed in the regional distribution of procedures from 2015 to 2023, as analyzed through linear regression.

**Table 5.** Procedures by region of the country.

| Region | SUS |  | PHS |  | Total |  |
| --- | --- | --- | --- | --- | --- | --- |
|  | *n* | % | *n* | % | *n* | % |
| North | 11,094 | 2.11% | 6,515 | 0.93% | 17,609 | 1.44% |
| Northeast | 55,745 | 10.58% | 73,904 | 10.56% | 129,649 | 10.57% |
| Midwest | 28,338 | 5.38% | 57,072 | 8.16% | 85,410 | 6.96% |
| Southeast | 309,058 | 58.66% | 398,613 | 56.97% | 707,671 | 57.70% |
| South | 122,619 | 23.27% | 163,592 | 23.38% | 286,211 | 23.33% |
| **Total** | 526,854 | 100.00% | 699,696 | 100.00% | 1,226,550 | 100.00% |

*SUS: Unified public health system; PHS: Private health system*

A total of 29 deaths were recorded following CVD surgery in the public healthcare system, corresponding to an overall mortality rate of 0.0056%. Although the Southeast region accounted for the highest absolute number of deaths, the North region exhibited the highest proportional mortality rate relative to the number of procedures performed (0.018%). Table 6 details mortality rates associated with CVD procedures in the public healthcare system. The D-TISS database does not report mortality associated with procedures performed in the PHS.

**Table 6.**
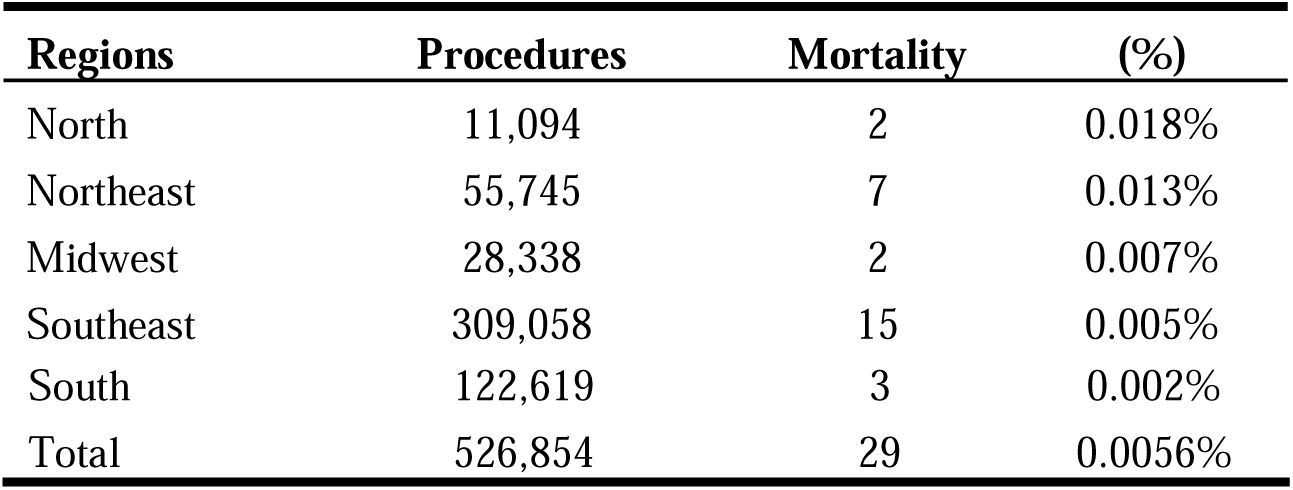
Absolute and relative distribution of deaths of patients undergoing surgical treatment for CVD in the Unified Health System (SUS).

## 7. Discussion

### Total Surgeries

A total of 1,266,550 surgeries were performed for the treatment of CVD. The public healthcare system accounted for 526,854 procedures, while the PHS performed 699,696 procedures. These interventions represent the largest series of vascular surgeries conducted during the evaluated period, highlighting the high prevalence of CVD and the ongoing need for these procedures to mitigate the effects of this condition on patient health. (22)

### Procedures rate per 10,000 inhabitants

The overall average rate was 6.5 CVD surgeries per 10,000 inhabitants annually in Brazil. Among exclusive SUS users, the rate was 3.7 per 10,000 inhabitants, while in the PHS, it was 14.8 per 10,000 inhabitants. For comparison, Japan reported an estimated rate of 4.14 per 10,000 (16), the United States 10.4 per 10,000 (23), England 12.1 per 10,000 (24), and Belgium 34.3 per 10,000, the highest reported in the literature. (25) These findings suggest that procedure rates in PHS are more comparable to those in developed countries than those in SUS.

When analyzing these disparities, it appears that the prevalence of CVD in Brazil and other countries is similar. (16,23,25) However, differences between SUS and European countries may be attributed to the smaller populations in European nations, which facilitates the management of low-complexity and low-mortality diseases such as CVD. Furthermore, public healthcare investment in Brazil is lower than in developed countries (26), which may also explain the discrepancies between SUS and PHS.

In terms of sex distribution, 77.65% of the recorded procedures were performed on women, while 21.50% were on men, and 0.085% of cases had unspecified sex. This finding is consistent with the well-documented higher prevalence of CVD in women compared to men. The proportions of male and female patients undergoing surgery were similar in both between SUS and PHS. (5,19,27)

Looking at age distribution, most procedures were conducted on adults aged 30 to 59, with the highest percentage occurring in the 40–49 age group (28.14%). While advanced age is a known risk factor for CVD and its complications, 18.47% of procedures were conducted in individuals over 60 years old. In a study analyzed 48,615 CVD surgeries in the United States, 21.3% of the procedures were performed on patients over 65 years, which is similar to our findings. (18)

Between 2015 and 2019, the total number of CVD surgeries in Brazil increased, predominantly due to the rise in procedures performed within PHS. However, a significant decline occurred in 2020 and 2021, coinciding with the COVID-19 pandemic. (28,29) This drop impacted the overall trend analysis for CVD surgeries. After this decline, the number of procedures rebounded in subsequent years, reaching pre-pandemic levels. In SUS, the peak year for procedures was 2018, while in PHS, the highest number of CVD surgeries occurred in 2022.

Regionally, the highest procedure rate was observed in the Southeast (57.70%), while the North region had the lowest rate (1.44%). Notably, the North was the only region where more procedures were performed within SUS than in PHS. However, this trend reversed after the pandemic, with PHS surpassing public healthcare. This shift may be attributed to an increase in the number of individuals enrolling in private health insurance plans during and after the pandemic. (30) Furthermore, all regions experienced a significant decline in procerures during 2020 and 2021, followed by a gradual recovery in subsequent years.

The variation in surgical procedure rates highlights socioeconomic inequality and differences in economic power and healthcare resources across Brazilian regions. (31) The Southeast, being the wealthiest and most populous region, had the highest number of procedures performed. In contrast, although the Northeast is the second most populous region, had less than half the number of procedures performed in the South, which has just over half of the Northeast’s population. (21)

These disparities are concerning because populations with lower socioeconomic status are at a higher risk for CVD and its complications, as well as other comorbidities that may complicate perioperative care. To improve (27,32)access to treatment in underserved regions, a range of interventions should be considered, from high-cost procedures procedures like radiofrequency ablation to lower-cost options such as foam sclerotherapy. (7,11,33,34)

### Government reimbursement - Supplementary health payments

Between 2015 and 2023, total expenditures for CVD treatment procedures in both public and private sectors reached R$ 1,492,310,372.13. The average annual cost per procedure across both systems was R$ 1,240.92. The public healthcare system allocated R$ 359,539,892.13, with an average expenditure of R$ 679.50 per procedure, while the PHS invested R$ 1,132,770,480.00, with an average of R$ 1,620.60 per procedure. This indicates that expenditures in PHS were 3.15-fold times higher than those in SUS, accounting for 76% of total spending during the analyzed period. Additionally, the number of procedures performed in PHS surpassed those in SUS by more than 100,000 cases. Despite SUS servicing approximately 75% of the Brazilian population, the 25% using PHS underwent nearly 60% of CVD surgeries, contributing to 76% of total expenditures.

International comparisons show that the per-procedure reimbursement in China and the U.K. is at least double the average amount allocated by SUS in Brazil and can be up to ten times higher in the United States. (33,35,36)

In the U.S., high costs are associated with ablative techniques performed in hospital settings. (33) However, due to data limitations, we were unable to differentiate distinguish between unilateral and bilateral procedures or the techniques used in SUS and PHS. Furthermore, SUS reimbursement follows a fixed compensation table, which may not reflect actual hospital costs, limiting deeper cost comparisons with other countries. In contrast, PHS expenditures are aligned with real case costs.

The disparity between the volume of procedures and healthcare costs in public versus private sectors is evident. Although Brazil provides universal healthcare, only 41% of total health expenditures come from public sources (37), which is relatively low compared to developed countries, where government health investments are significantly higher. (26) This funding gap impacts CVD treatment, as PHS financed 76% of surgical costs despite serving only 25% of the population.

### Deaths from the Public Health System

Among the 1,266,550 CVD surgeries performed, mortality data were only available for cases reported in the public healthcare system (526,584 cases). A total of 29 deaths were recorded during hospitalization or in outpatient clinics, resulting in a mortality rate of 0.005%. A 2022 epidemiological analysis of CVD cases in the Brazilian public healthcare system from 2008 to 2019 reported a similar mortality rate. (19) In comparison, the in-hospital mortality rate in Portugal was 0.015%, which is approximately three times higher than in Brazil. (17) In the United States, a study on elderly patients showed a 30-day mortality rate of 0.02% (18), while in astudy conducted in Japan involving 52,639 cases reported no deaths within 30 days post-surgery. (16) These findings highlight the low mortality risk associated with CVD surgery, even among higher-risk populations.

This study has limitations regarding mortality assessment. Due to data anonymization on the DATASUS platform, the causes of death were not identified, and patient records were not analyzed, which prevented the evaluation of deaths occurring after hospital discharge. Consequently, 30-day mortality could not be calculated. This may lead to underreporting of fatal but non-immediate complications, such as pulmonary embolism, while perioperative deaths from anesthesia or hemorrhagic complications were more likely to be captured. Additionally, the absence of mortality data in the D-TISS platform further underestimates the actual number of deaths.

### Limitations

A retrospective or longitudinal analysis of operated patients was not feasible, as data from both DATASUS and D-TISS are anonymized and provide only the total number of procedures. Consequently, the exact number of treated patients remains unknown, as some may have undergone multiple procedures or reinterventions. Additionally, this study is limited by the potential misclassification and underreporting of cases.

Despite these limitations, this study presents a comprehensive analysis of all CVD surgical procedures conducted nationwide (both in SUS and PHS) between 2015 and 2023. It offers real-world data with extensive external validation, highlighting significant differences in treatment volume and costs between the two healthcare systems.

## 8. Conclusion

Between 2015 and 2023, a total of 1,266,550 CVD surgeries were performed , resulting in an annual rate of 6.5 procedures per 10,000 inhabitants. The number of procedures conducted in the PHS was four times higher than those in SUS when considering the rate per 10,000 inhabitants. The total investment in PHS was 3.15 times higher than in SUS, with an estimated average cost per procedure of R$ 1,632.60. The observed mortality rate in SUS was 0.005%.

## Research Ethics Approval

This study was approved by the Ethics Committee of the **institution** where it was carried out.

## Data Availability

All data produced in the present study are available upon reasonable request to the authors

